# Multi-centre discovery and validation study evaluating breath biomarkers for the diagnosis of lung cancer – the LuCID study

**DOI:** 10.1101/2025.06.29.25330417

**Authors:** Marc van der Schee, Max Allsworth, Yichen Chen, Rob Smith, Simon Kitchen, Mariana Ferreira Leal, Chris Hodkinson, Billy Boyle, Alexandra de Saedeleer, Duncan Apthorp, Silvano Dragonieri, Hubert Wirtz, Anjani Prasad, Mohammed Haris, Liz Fuller, Jacqueline Faccenda, David R Baldwin, Arnaud Scherpereel, Jonathan Bennett, Serena Chee, Andrew Barlow, Andrew Wight, Martin Ledson, Eleanor Mishra, William Ricketts, Seamus Grundy, Philip Crosbie, Mamta Ruparel, Samuel M Janes, Robert C Rintoul

## Abstract

**Background:** Volatile Organic Compound (VOC) research for lung cancer detection has faced study design and analytical methodology challenges limiting translation into clinical practice. We evaluated the diagnostic value of breath biomarkers in patients under investigation for suspected lung cancer.

**Methods:** In a multi-centre prospective case-control study involving 1844 subjects under investigation, breath samples from subjects with a conclusive diagnosis were analysed using gas-chromatography mass-spectrometry. A staged approach was adopted: an Exploratory method for targeted analysis of 63 VOCs associated with lung cancer, followed by an Optimised method for biomarker discovery and finally, evaluation of the optimised panel in a separate validation cohort. Results were compared to the Liverpool Lung Project (LLP) risk model.

**Findings:** Using breath VOCs from 677 controls and 518 cases the Exploratory method showed only 2 literature-reported compounds differed significantly between cases and controls. The Optimised method detected 102 VOCs, with ten differing between cases and controls. However, in a validation cohort the 10-VOC panel differentiated cases from controls cohort with a modest AUC: 0.54±0.14 for early-stage disease, 0.58±0.16 for advanced stage disease and 0.58±0.11 for all cases, which did not differ significantly from the LLP model. Combining VOCs with the LLP model did not significantly improve diagnostic performance (AUC 0.64±0.11).

**Interpretation:** Although some potential biomarkers were identified, their diagnostic performance did not surpass an epidemiological risk model. The study highlights the importance of careful trial design to avoid false-positive findings and indicates a need for more targeted approaches to enhance signal-to-noise ratio in breath biomarker research.

**Highlights:** Largest, to date, multi-centre prospective case-control study evaluating volatile organic compounds (VOC) for intention-to-diagnose lung cancer.

Gas chromatography mass spectrometry was used to compare VOCs in lung cancer cases with co-morbidity matched controls.

Although some VOCs differentiated cases from controls, diagnostic performance did not surpass an epidemiological risk model.

These data explain the challenge of previous studies to validate and translate into clinical practice.

A more targeted approach to enhance signal-to-noise ratio is required.

## Introduction

The premise of breath analysis for the early detection of cancer is its ability to reflect metabolic changes inside the body by analysing Volatile Organic Compounds (VOCs) that occur in trace amounts in the breath^1^. As metabolic changes occur early in the development of cancer^2^, VOCs may have advantages for the early detection of cancer over other approaches such as circulating tumour DNA which, to date, is limited in its sensitivity for detection of the earliest stages of disease^3^.

In the earlier detection of lung cancer field, Low Dose Computed Tomography (LDCT) has been shown to decrease mortality from lung cancer^4,5^. However, implementation challenges around identifying optimal target populations and combatting low screening rates among high-risk individuals persist^6,7^. To enhance LDCT adoption, the Lung Cancer Screening Policy Network recommends tailored eligibility criteria, targeted outreach, and biomarker-based prioritization to improve cost-efficiency and encourage adoption in harder-to-reach populations^8^. In this context non-invasive breath analysis has received considerable attention to drive adoption and accessibility of lung cancer screening^9^. However, despite considerable research on VOCs as potential biomarkers, there is a notable discrepancy between the extensive scientific investigations and translation into clinical practice^9^. This gap is rooted in several challenges such as issues related to study design, analytical methodology and statistical approaches (Supplementary Table S1). At present, the potential of breath biomarkers for detection of lung cancer, including lung cancer in a screening setting, remains unclear.

Here, we describe the results of the LuCID (Lung Cancer Indicator Detection) study which was designed to address the aforementioned challenges and provide a high degree of confidence for the evaluation of breath biomarkers for lung cancer using state-of-the-art breath collection and analysis approaches. This study evaluated breath biomarkers collected at the time of clinical presentation using targeted and untargeted gold standard Mass-Spectrometry techniques to identify biomarkers that distinguish between individuals with and without lung cancer. This approach allowed investigation of both previously reported breath biomarkers for lung cancer and the exploration of novel biomarkers. To mimic clinical implementation, the diagnostic value of breath biomarkers was assessed by evaluating their (additive) performance relative to a clinical risk score (Liverpool Lung Project risk score^10^). As such, the LuCID study provides detailed insights into the value of naturally occurring breath VOCs as a tool for the (earlier) detection of lung cancer.

## Methods

### Design

LuCID was a prospective case-control cohort study enrolling consecutive individuals suspected of having lung cancer from 26 study sites across the United Kingdom and European Union, between October 2015 and March 2020 (see Supplementary method for study sites). The primary endpoint was the evaluation of the diagnostic accuracy of breath biomarkers for the detection of lung cancer stratified by early-stage and advanced-stage disease. Relevant breath biomarkers were evaluated using two breath analysis methods in two independent cohorts: 1) Exploratory method, 2) Optimised method.

Using the Exploratory method, breath collection and analysis approach were optimized for the detection of previously reported breath biomarkers for lung cancer. In addition, the Exploratory method was developed with the capability to identify new candidate biomarkers. Interim results were utilised to optimise the breath collection and analysis methodology. Breath samples from the second prospectively recruited cohort were analysed using an Optimised methodology with an improved signal-to-noise ratio thereby allowing identification of breath VOCs not identified by the Exploratory method. For both Exploratory and Optimised approaches, subjects were randomly assigned to training (75%) and validation sets (25%). Whenever possible, the relevance of individual VOCs was assessed by determining their chemical identity through structural elucidation, to allow biochemical linkage to cancer pathobiology.

### Participants

Eligible individuals were referred for investigation with symptoms, or imaging, suspicious for lung cancer. The study excluded subjects on active treatment for any type of malignancy. Following signed informed consent, relevant data on demographics, smoking history, comorbidities, medications, occupational exposures and relevant family history were collected. The Liverpool Lung Project v2.0 score was calculated for each participant^10^. All study subjects provided a breath sample using the ReCIVA Breath Collector (Owlstone Medical, Cambridge, UK) prior to the completion of their diagnostic work-up. Lung cancer cases were defined as those with a pathologically confirmed diagnosis of lung cancer based on an expert discussion in a multidisciplinary team meeting. Lung cancer stages Ia-IIb (7^th^ TNM edition) were considered early-stage lung cancer, while stages III-IV were classified as advanced-stage disease. Controls subjects consisted of patients initially presenting with symptoms and/or imaging suspicious for lung cancer through the same pathways as the lung cancer cases, but with a final diagnosis of a benign condition. Cases diagnosed with non-pulmonary cancers and controls diagnosed with any cancer at any time during a 24-month follow-up period after breath sampling were excluded. Participants with an inconclusive diagnostic work-up, most commonly indeterminate pulmonary nodules or absence of histology, were also excluded. To ensure data consistency, all diagnostic data were independently monitored. Full inclusion and exclusion criteria are listed in Supplementary methods.

### Breath sample collection

Breath samples were collected using the ReCIVA® breath collector developed as part of the open-source Breath-Free consortium^11^ in response to European Respiratory Society taskforce on existing challenges with breath analysis^12^. The device enables selective sampling of breath fractions via pressure gating. Each breath sample was collected onto four sorbent tubes (Tenax TA, Carbograph 5TD, Markes International, Bridgend, UK), optimized for hydrophobicity and capture of a wide range of VOCs. A total volume of 0.5L of alveolar enriched breath was collected for the Exploratory method and 1.5L of full exhaled breath for the Optimised method. Quality control was performed through algorithmic analysis of the pressure sensors in ReCIVA® which ensured adequate collection quality and volume. Both methods integrated the use of the CASPER^TM^ clean air supply (Owlstone Medical, Cambridge, UK), which provides low-VOC inhalation air to reduce contribution of environmental background VOCs into breath samples. Collected samples were sealed and stored in a fridge and shipped to Owlstone Medical within 7 days.

### Breath VOCs analysis

Upon receipt, samples were either stored at 4°C (Exploratory method) or purged with helium to remove water prior to storage at −20°C (Optimized method). Samples were analysed by thermal desorption-gas chromatography-mass spectrometry (TD-GC-MS, TD-100-XR Markes Int., Trace GC 1300 Thermo scientific, Bench TOF Markes Int., Bridgend, UK). The Exploratory method utilised a general-purpose approach to cover a broad range of 63 literature reported analytes (Supplementary Table S2). Samples were desorbed onto a non-polar column for a 60-minute analysis with 35 minutes analytical space. The Optimized method aimed at improving analytical space and increasing the signal of VOCs originating from the study subject whilst reducing chemical background. Samples were desorbed splitless onto a mid-polarity column with 86.5 minutes run time and 71 minutes analytical space. Chromatographic resolution was monitored using standards. Mass spectral intensity was controlled via tune function and adjusting source parameters – samples were not analysed until the system was demonstrated to be within expected performance. Detailed descriptions of the methodology used are available in the supplementary material. To address the well-established challenge of feature alignment in -omics studies^13^, both targeted and untargeted approaches were employed for feature extraction. Chemical standards were run for both methods to obtain reference spectra, ensuring reliable and consistent identification of reported compounds. Additional biomarker discovery was conducted by extracting unknown features based on their relative positions to known targets from standards. These unknown VOC features were then elucidated by generating a list of possible candidates using National Institute of Standards and Technology database matching, refined by comparing the expected retention time between candidate and feature, and, where possible, confirmed by running standards alongside replicate breath samples. Not all features were successfully elucidated utilising state of the art elucidation strategies, resulting in labelling a feature as unknown. Further detail on analytical methodology and feature extraction approaches is provided in the supplementary materials.

### Data-analysis

Breath VOC data were processed separately for each cohort (Exploratory and Optimized methods) independent of clinical data. Missing values in VOC intensities were imputed to 80% of the minimum observed intensity for each VOC. Instrument sensitivity drift over time and between instruments was normalized for each VOC. VOCs underwent log-transformation before statistical analysis to mitigate the impact of samples with high intensities. Variables were standardized before modelling to improve numerical stability. For both the Exploratory and Optimised methods, subjects were randomly assigned to training (75%) and validation sets (25%), ensuring balanced histological types. The hold-out set was used to determine diagnostic accuracy. Further detail on data analysis is provided in supplementary materials.

Univariate analysis identified VOCs associated with the presence of lung cancer. The training set was analysed using a linear mixed-effects model with VOC intensity as the outcome, case-control status as the predictor, and collection site as the random intercept to control for site bias. This analysis was conducted by adjusting for age, gender, BMI, and smoking status as model covariates. The Benjamini-Hochberg procedure was applied to correct for multiple testing, with a P-value < 0.05 considered statistically significant.

Using data generated with the Optimized method, multivariate logistic regression was applied to generate a classifier. The predictors included VOCs alone, and in combination, with the Liverpool Lung Project risk model score. VOCs with P-values < 0.05 in both covariate-adjusted and unadjusted univariate analyses were used as candidate predictors to avoid overfitting. Elastic-net regularization (L1 ratio=0.9) aided feature selection. A 20-split Monte Carlo cross-validation applied to the training set determined the optimal regularization strength. The final model featured consistently selected variables across 90% of splits. Validation set performance was assessed using the ROC-AUC, with 95% confidence intervals. DeLong’s test was used to compare the AUCs. All univariate and multivariate analyses were conducted using all lung cancer cases (free from other cancers) and then repeated separately for early-stage and advanced-stage lung cancer.

Power calculation was based on within-class and between-site variation from the current study by computing the power of a similarly sized study in detecting a single VOC that would have a hypothetical case-control difference equivalent to 60% within-site specificity and sensitivity. The power calculation assumed absence of class imbalance and was based on a single-covariate linear mixed-effect model with a p-value cut-off at 0.05. Further detail is provided in supplementary materials.

### Role of the funding source

Owlstone Medical Ltd sponsored the study. Study design, data interpretation and writing of the manuscript was a collaboration between the academic authors and Owlstone Medical Ltd. Breath samples and patient data were collected by the academic investigators in clinic settings and sample processing and VOC analyses were performed by Owlstone Medical (Cambridge, UK) who was blinded to patient’s characteristics and outcomes. The decision to submit the paper for publication was taken by all authors.

### Study governance

The study received ethical approval from the NRES Committee East of England – Cambridge South in October 2015 (15/EE/0298). The study was reported according to the Standards for Reporting of Diagnostic Accuracy Studies (STARD) reporting guidelines.

LuCID was registered ClinicalTrials.gov NCT02612532.

## Results

A total of 1,844 subjects were recruited (Figure 1). Of these, 342 (18.5%) were excluded from the final analyses after clinical curation: 336 because they lacked unequivocal per-protocol tissue-based diagnoses (primarily due to indeterminate pulmonary nodules or unsuccessful biopsies), and 6 participants who met exclusion criteria through clinical data monitoring. One hundred fourteen participants (6.2% of the recruited subjects) were excluded due to technical issues: 67 for technical failure of breath collections and 47 due to analytical failures. Additionally, 193 subjects were diagnosed with non-pulmonary malignancies (56 cases) or any malignancy (137 controls) during the two-year follow-up and were excluded from the per-protocol analysis. Therefore, breath VOC data were obtained for 677 controls and 518 cases. Breath biomarker data for a total of 809 samples analysed using the Exploratory method and an additional 386 using the Optimised method were used as input for the data analysis. Results presented below are stratified by analytical methodology (Exploratory or Optimised) and provided for all stages of lung cancer, further stratified into early-stage (I and II) and advanced-stage (III and IV). Supplementary Table 3 provides details on the results of the clinical work-up for participants who were found not to have lung cancer following diagnostic work-up. Where known, many of these were infectious/inflammatory aetiologies including infectious exacerbations of Chronic Obstructive Pulmonary Disease (COPD) or interstitial lung disease. There were no serious adverse events from collection of breath samples.

**Figure 1.**
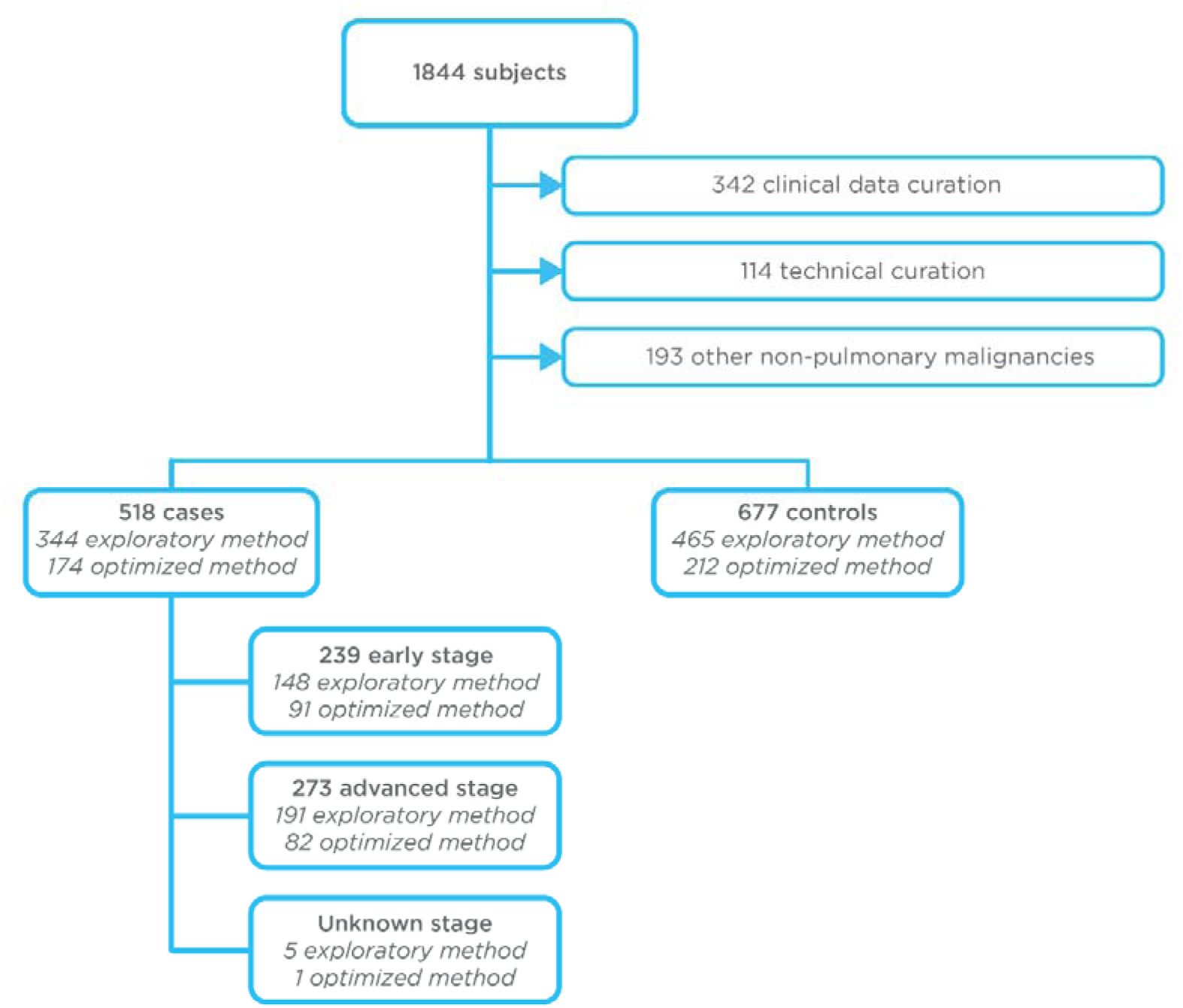
Consort diagram for the LUCID study. Primary analyses compared a cumulative of 677 controls and 518 cases across the *Exploratory* and *Optimised* analytical methods. Clinical and technical curation was performed to exclude subjects with unresolved diagnostic work-up (i.e. indeterminate nodules) or erroneous breath collections or analyses. Early Stage: Stage I and II; Advanced Stage: Stage III and IV. * Five subjects recruited as part of the *Exploratory* method and 1 subject recruited as part of the *Optimised* method had a histologically confirmed tumour but unconfirmed tumour stage. These samples were included in the “all-stages” analysis but excluded from the stratified analysis by tumour stage. For both the *Exploratory* and *Optimised* methods, subjects were randomly assigned to training (75%) and validation sets (25%), ensuring balanced histological types.

### Exploratory method

A total of 809 breath samples were analysed using the Exploratory analytical method: 465 from controls and 344 from cases (148 with early-stage lung cancer, 191 with late-stage lung cancer, and 5 with undetermined stage). As expected, in the intention-to-diagnose population, known risk factors such as age, years smoked, COPD and family history of lung cancer in first degree relative were significantly different between cases and controls (p < 0.05; Table 1a).

**Table 1.**
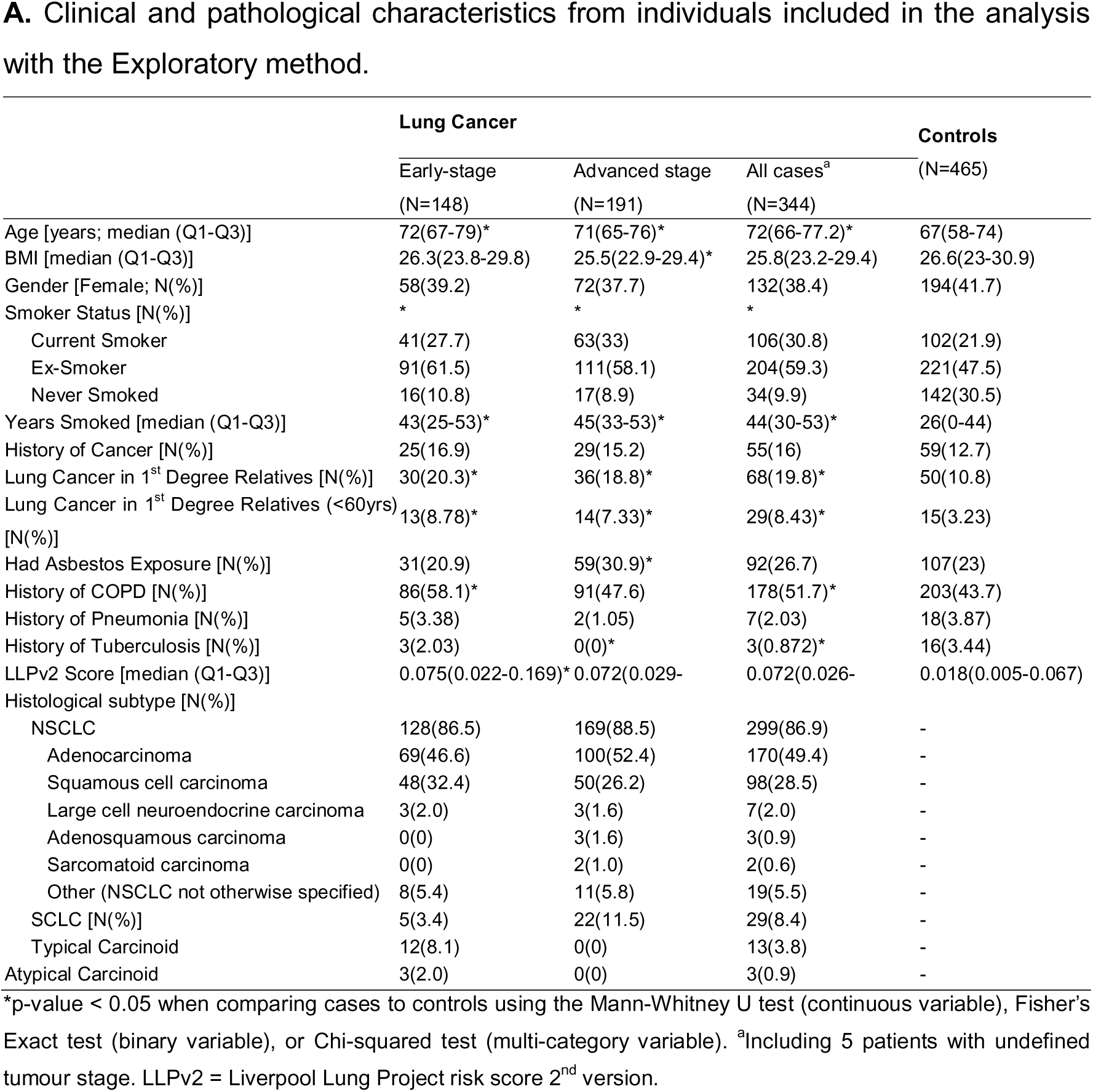
Clinical and pathological characteristics from individuals included in the analysis with the Exploratory method.

|  | Lung Cancer |  |  | Controls<br>(N=465) |
| --- | --- | --- | --- | --- |
|  | Early-stage<br>(N=148) | Advanced stage<br>(N=191) | All cases<sup>a</sup><br>(N=344) |  |
| Age [years; median (Q1-Q3)] | 72(67-79)* | 71(65-76)* | 72(66-77.2)* | 67(58-74) |
| BMI [median (Q1-Q3)] | 26.3(23.8-29.8) | 25.5(22.9-29.4)* | 25.8(23.2-29.4) | 26.6(23-30.9) |
| Gender [Female; N(%)] | 58(39.2) | 72(37.7) | 132(38.4) | 194(41.7) |
| Smoker Status [N(%)] | * | * | * |  |
| Current Smoker | 41(27.7) | 63(33) | 106(30.8) | 102(21.9) |
| Ex-Smoker | 91(61.5) | 111(58.1) | 204(59.3) | 221(47.5) |
| Never Smoked | 16(10.8) | 17(8.9) | 34(9.9) | 142(30.5) |
| Years Smoked [median (Q1-Q3)] | 43(25-53)* | 45(33-53)* | 44(30-53)* | 26(0-44) |
| History of Cancer [N(%)] | 25(16.9) | 29(15.2) | 55(16) | 59(12.7) |
| Lung Cancer in 1<sup>st</sup> Degree Relatives [N(%)] | 30(20.3)* | 36(18.8)* | 68(19.8)* | 50(10.8) |
| Lung Cancer in 1<sup>st</sup> Degree Relatives (<60yrs)<br>[N(%)] | 13(8.78)* | 14(7.33)* | 29(8.43)* | 15(3.23) |
| Had Asbestos Exposure [N(%)] | 31(20.9) | 59(30.9)* | 92(26.7) | 107(23) |
| History of COPD [N(%)] | 86(58.1)* | 91(47.6) | 178(51.7)* | 203(43.7) |
| History of Pneumonia [N(%)] | 5(3.38) | 2(1.05) | 7(2.03) | 18(3.87) |
| History of Tuberculosis [N(%)] | 3(2.03) | 0(0)* | 3(0.872)* | 16(3.44) |
| LLPv2 Score [median (Q1-Q3)] | 0.075(0.022-0.169)* | 0.072(0.029- | 0.072(0.026- | 0.018(0.005-0.067) |
| Histological subtype [N(%)] |  |  |  |  |
| NSCLC | 128(86.5) | 169(88.5) | 299(86.9) | - |
| Adenocarcinoma | 69(46.6) | 100(52.4) | 170(49.4) | - |
| Squamous cell carcinoma | 48(32.4) | 50(26.2) | 98(28.5) | - |
| Large cell neuroendocrine carcinoma | 3(2.0) | 3(1.6) | 7(2.0) | - |
| Adenosquamous carcinoma | 0(0) | 3(1.6) | 3(0.9) | - |
| Sarcomatoid carcinoma | 0(0) | 2(1.0) | 2(0.6) | - |
| Other (NSCLC not otherwise specified) | 8(5.4) | 11(5.8) | 19(5.5) | - |
| SCLC [N(%)] | 5(3.4) | 22(11.5) | 29(8.4) | - |
| Typical Carcinoid | 12(8.1) | 0(0) | 13(3.8) | - |
| Atypical Carcinoid | 3(2.0) | 0(0) | 3(0.9) | - |

\*p-value < 0.05 when comparing cases to controls using the Mann-Whitney U test (continuous variable), Fisher's Exact test (binary variable), or Chi-squared test (multi-category variable). <sup>a</sup>Including 5 patients with undefined tumour stage. LLPv2 = Liverpool Lung Project risk score 2<sup>nd</sup> version.

**Table 1.**
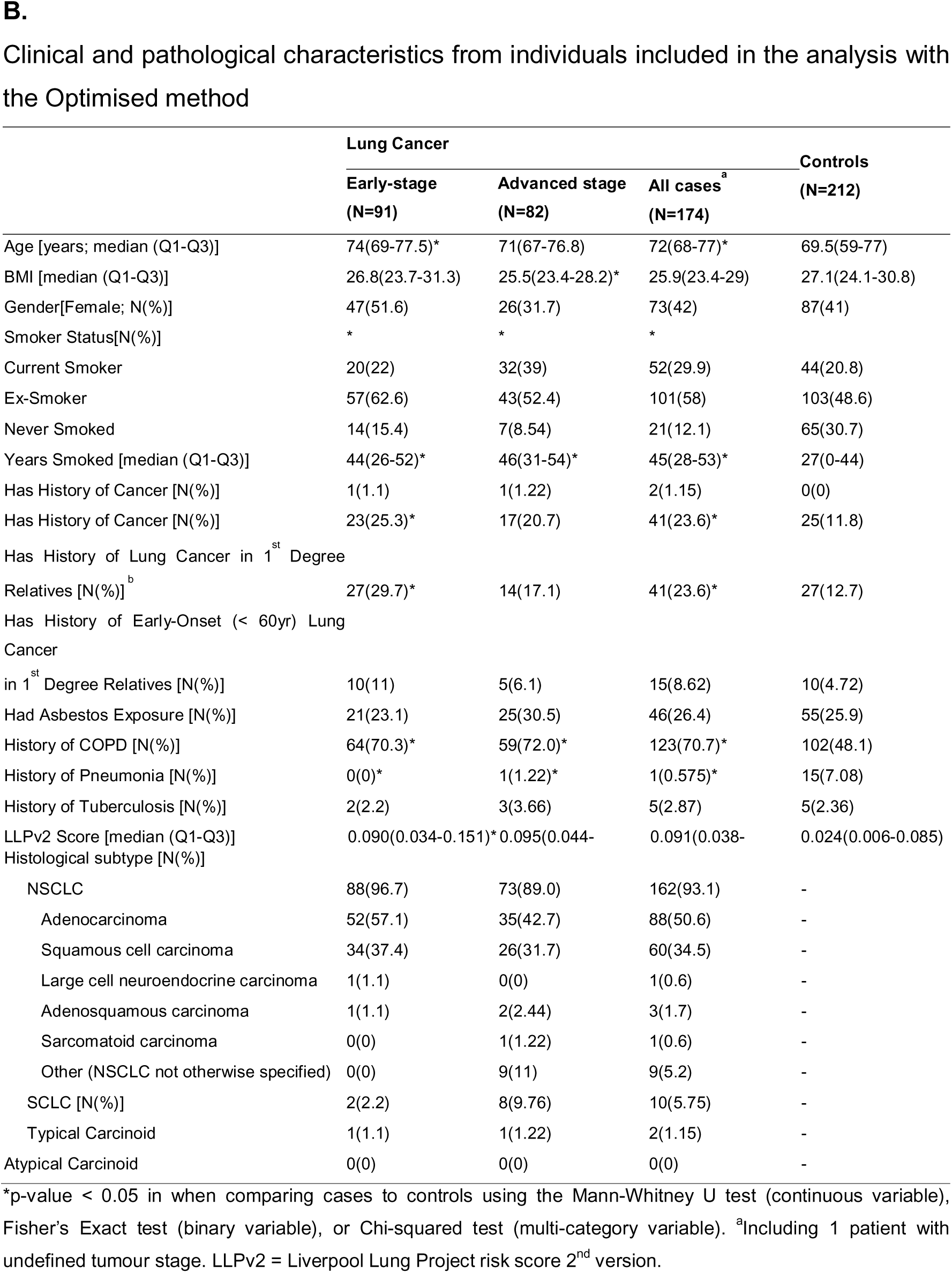
Clinical and pathological characteristics from individuals included in the analysis with the Optimised method.

In total, 84 potential VOCs of interest were extracted from the spectral data, including 49 of 63 from the VOC target panel (Supplementary Table S4). Analysis, uncorrected for multiple testing, was used to identify potential candidate markers.

This analysis identified four compounds as specified in Table 2a. Allyl methyl sulphide and acetoin were lower in subjects with lung cancer compared to controls, an association driven by advanced cancer stages (Figure 2A and 2C). D-limonene showed a similar trend (Table 2a), with p < 0.05 but only when comparing advanced cancer to controls. On the other hand, 2-undecanone appeared in the breath of advanced lung cancer subjects at higher levels than in controls. Of the four potential candidate markers identified in the Exploratory analysis, D-limonene^15^ and 2-undecanone^16^ were previously reported in the literature and featured in the 63 VOC target panel whereas Allyl Methyl Sulphide and Acetoin were potential new candidate biomarkers. None of the remaining compounds in the target panel were either detectable or associated with lung cancer in this population. However, following correction of p-values for multiple comparisons, none of the four candidate VOC biomarkers were associated with the presence of lung cancer in either covariate-adjusted or unadjusted analyses. Therefore, diagnostic accuracy analysis on the 25% validation cohort was not performed. A full breakdown of the analysis per target analyte is provided in Supplementary Table S4. The power for the Exploratory method in detecting a single VOC that has a hypothetical within-site effect size equivalent to 60% specificity and 60% sensitivity was at least 99.6%.

**Table 2.** Univariate VOC analysis in the cohort evaluated with the Exploratory method.

A. Univariate VOC analysis in the cohort evaluated with the Exploratory method.
| Breath<br>(VOC) | biomarker | Co-variate<br>correction applied | Early-stage |  | Advanced stage |  | All cases |  |
| --- | --- | --- | --- | --- | --- | --- | --- | --- |
|  |  |  | Coefficient | p-value | Coefficient | P-value | Coefficient | p-value |
| Allyl Methyl Sulphide |  | Uncorrected | -0.254 | 0.0183 | -0.386 | <0.001 | -0.329 | <0.001 |
|  |  | Corrected | -0.124 | 0.265 | -0.263 | 0.011 | -0.218 | 0.011 |
| Acetoin |  | Uncorrected | -0.104 | 0.348 | -0.344 | 0.001 | -0.251 | 0.002 |
|  |  | Corrected | -0.075 | 0.512 | 0.250 | 0.016 | -0.209 | 0.015 |
| D-Limonene |  | Uncorrected | 0.117 | 0.279 | -0.287 | 0.004 | -0.111 | 0.187 |
|  |  | Corrected | 0.126 | 0.262 | -0.314 | 0.003 | -0.131 | 0.143 |
| 2-Undecanone |  | Uncorrected | 0.111 | 0.326 | 0.229 | 0.031 | 0.194 | 0.026 |
|  |  | Corrected | 0.114 | 0.337 | 0.273 | 0.015 | 0.228 | 0.014 |

B. Univariate VOC analysis in the cohort evaluated with the Optimised method.
| Breath<br>(VOC) | biomarker | Co-variate<br>correction applied | Early-stage |  | Advanced stage |  | All cases |  |
| --- | --- | --- | --- | --- | --- | --- | --- | --- |
|  |  |  | Coefficient | p-value | Coefficient | P-value | Coefficient | p-value |
| cyclohexane |  | Uncorrected | -0.123 | 0.474 | -0.329 | 0.036 | -0.213 | 0.081 |
|  |  | Corrected | -0.128 | 0.494 | -0.357 | 0.034 | -0.21 | 0.122 |
| Allyl methyl sulphide |  | Uncorrected | -0.182 | 0.33 | -0.571 | <0.001 | -0.406 | 0.004 |
|  |  | Corrected | -0.048 | 0.762 | -0.444 | 0.01 | -0.319 | 0.032 |
| Acetoin |  | Uncorrected | -0.188 | 0.349 | -0.397 | 0.012 | -0.317 | 0.009 |
|  |  | Corrected | -0.203 | 0.332 | -0.454 | 0.005 | -0.327 | 0.012 |
| Acetophenone |  | Uncorrected | -0.356 | 0.02 | -0.177 | 0.274 | -0.263 | 0.056 |
|  |  | Corrected | -0.344 | 0.033 | -0.215 | 0.219 | -0.295 | 0.047 |
| unk12 |  | Uncorrected | -0.221 | 0.151 | 0.345 | 0.022 | 0.063 | 0.607 |
|  |  | Corrected | -0.292 | 0.072 | 0.372 | 0.023 | 0.038 | 0.774 |
| unk45 |  | Uncorrected | -0.323 | 0.035 | -0.121 | 0.427 | -0.217 | 0.076 |
|  |  | Corrected | -0.346 | 0.033 | -0.049 | 0.762 | -0.201 | 0.122 |
| Geranylacetone<br>(cis/trans) |  | Uncorrected | -0.382 | 0.012 | -0.201 | 0.187 | -0.274 | 0.025 |
|  |  | Corrected | -0.462 | 0.004 | -0.245 | 0.134 | -0.329 | 0.011 |
| Hexadecane |  | Uncorrected | -0.441 | 0.026 | -0.193 | 0.236 | -0.228 | 0.105 |
|  |  | Corrected | -0.515 | 0.013 | 0.140 | 0.421 | -0.235 | 0.116 |
| (1-ethylnonyl)-<br>benzene (tentative) |  | Uncorrected | -0.329 | 0.032 | -0.194 | 0.226 | -0.268 | 0.028 |
|  |  | Corrected | -0.368 | 0.023 | -0.258 | 0.126 | -0.305 | 0.018 |
| unk23 |  | Uncorrected | -0.502 | 0.001 | -0.25 | 0.114 | -0.364 | 0.003 |
|  |  | Corrected | -0.517 | 0.001 | -0.321 | 0.057 | -0.389 | 0.002 |

Analysis uncorrected for multiple testing identified 10 VOCs as potential breath biomarkers for lung cancer. Upon applying multiple-testing correction this association was demonstrated to be non-significant.

**Figure 2.**
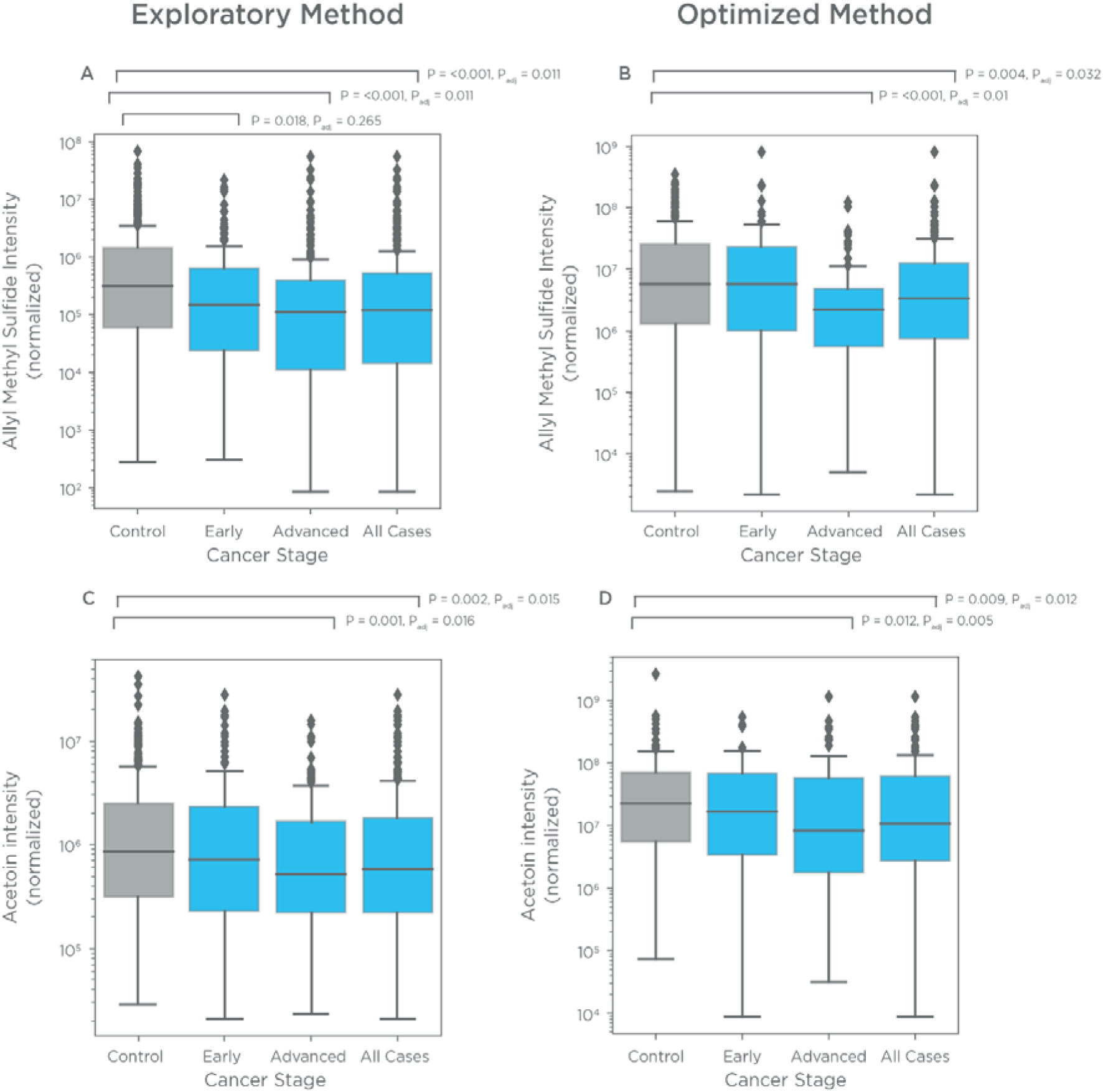
Normalized levels of volatile organic compounds on breath. A) Allyl Methyl Sulphide in cohort evaluated with *Exploratory* method; B) Acetoin in cohort evaluated with *Exploratory* method; C) Allyl Methyl Sulphide in cohort evaluated with *Optimised* method; D) Acetoin in cohort evaluated with *Optimised* method. P-values (prior to multiple testing correction) before and after covariate adjustment are displayed.

### Optimised method

Data from the Exploratory method were used alongside the refined collection and analysis approaches of the Optimised method to enhance biomarker discovery. A total of 386 breath samples were analysed using the Optimised methodology, including 212 controls and 174 cases (91 with early-stage lung cancer, 82 with late-stage lung cancer, and 1 with an undetermined stage). Subject characteristics are detailed in Table 1b. In total, 102 potential VOCs of interest were extracted from the spectral data, including 51 out of 63 from the VOC target panel (see supplementary table S5).

Patients with lung cancer (early or advanced disease) had significantly lower levels of ten VOCs compared to controls (p < 0.05 by both covariate-adjusted and unadjusted univariate analysis; Table 2b). The VOCs acetophenone, hexadecane, unknown VOC 45, geranyl acetone (cis or trans), (1-ethylnonyl)-benzene (tentative), and unknown VOC 23 (undetermined terpenoid) were significantly lower in the breath of patients with early-stage lung cancer compared to controls. Conversely, cyclohexane, allyl methyl sulphide (Figure 2B), and acetoin (Figure 2D) were lower in breath samples from patients with advanced-stage lung cancer compared to controls. In addition, patients with advanced lung cancer had high levels of one VOC in breath (unknown VOC 12) compared to controls (p < 0.05 by both covariate-adjusted and unadjusted univariate analysis; Table 2b). Of these compounds, 4 have previously been reported in the literature and were part of the target analyte list. As observed in the cohort evaluated with the exploratory method, none of these differences passed multiple testing correction. A full breakdown of the analysis is provided in Supplementary Tables S5. The power for the Optimised method in detecting a single VOC that has a hypothetical within-site effect size equivalent to 60% specificity and 60% sensitivity was at least 88.2%.

### Multivariate diagnostic classifier

The ten VOCs identified as differing between cases and controls in the Optimised method were used as input for developing a multivariate diagnostic classifier. However, the diagnostic accuracy in the 25% balanced validation cohort for differentiating between controls and cases with early-stage lung cancer (AUC: 0.54 ± 0.14), advanced-stage lung cancer (AUC: 0.58 ± 0.16), and all lung cancer cases (AUC: 0.58 ± 0.11) was poor (Figure 3; Table 3). No significant difference in AUC was observed between the Liverpool lung epidemiological risk model v2.0 (AUC 0.66 ± 0.11) and the VOC-only model (Figure 3; Table 3). Furthermore, the combination of VOCs with the Liverpool Lung Risk model did not significantly improve diagnostic performance (AUC 0.64 ± 0.11).

**Figure 3.**
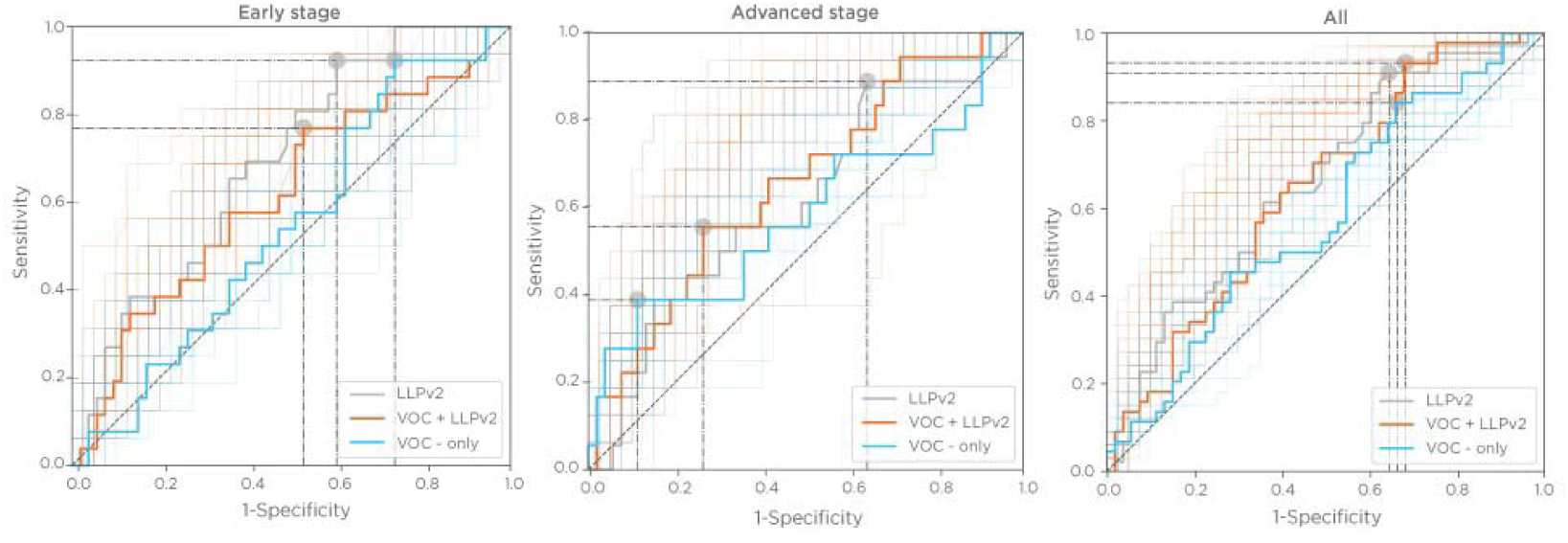
ROC-Curves for multivariate classification models. ROC-Curves for the 3 reported models: VOC only (blue colour), Liverpool Lung Risk Model (grey colour) and VOC & Liverpool Lung Risk Model (orange colour) in the Optimised cohort composed of controls and early-stage lung cancer, advanced stage lung cancer and all patients with lung cancer. Empirical variation in ROC curve was simulated by repeating the model-building and the ROC construction process via a 20-split Monte-Carlo cross-validation applied to the training set.

**Table 3.** Diagnostic performance for multivariate analysis with and without breath biomarkers in the cohort evaluated with the Optimised method.

| Stage | Model | ROC-AUC<br>(95%CI) | Sensitivity (%) | Specificity<br>(%) | Positive<br>Predictive Value<br>(PPV) (%) | Negative Predictive<br>Value (NPV) (%) |
| --- | --- | --- | --- | --- | --- | --- |
| Early-stage | VOC only | 0.543(±0.137) | 92.3 | 26.4 | 37.1 | 82.4 |
|  | LLPv2 | 0.689(±0.130) | 92.3 | 39.6 | 41.8 | 87.5 |
|  | VOC & LLP | 0.615(±0.135) | 76.9 | 47.2 | 40.4 | 78.1 |
| Advanced stage | VOC only | 0.584(±0.157) | 38.9 | 88.7 | 50.0 | 79.7 |
|  | LLPv2 | 0.615(±0.156) | 88.9 | 35.8 | 31.3 | 87.0 |
|  | VOC & LLP | 0.654(±0.154) | 55.6 | 73.6 | 39.1 | 81.3 |
| All Cases | VOC only | 0.580(±0.115) | 84.1 | 34.0 | 50.7 | 69.2 |
|  | LLPv2 | 0.659(±0.110) | 90.9 | 35.8 | 54.2 | 80.0 |
|  | VOC & LLP | 0.642(±0.111) | 93.2 | 32.1 | 52.6 | 81.0 |

Values are reported at the Youden Index of the ROC-Curve (best overall performance). Values represent a breath biomarker only (VOC-only), Liverpool Lung Risk Project (LLPv2) based model and a combined VOC & LLPv2 model.

## Discussion

The LuCID study demonstrated that the ability of a state-of-the-art GC-MS breath biomarker discovery approach to diagnose lung cancer was poor and no more accurate than an epidemiological risk model. The study identified several consistent associations of VOCs with the presence of lung cancer. However, while these associations appear robust, their variability within case and control groups is substantial and outweighs the differences between these groups. The study also highlighted the significant impact that multiple testing correction and accounting for covariates such as age and smoking, have on the results. In conclusion, while some modest diagnostic capability exists, the performance of a VOC-based diagnostic classifier is comparable to clinically available risk factors.

These results contrast with many breath analysis studies in lung cancer in terms of their outcomes^9^. Across the two analytical methods utilized in this study, a combined total of 58 of 63 (92%) VOCs previously reported in the literature were successfully identified, with 42 of 58 in common between the two methods. However, only around 10% of these 58 showed any diagnostic potential across the cohorts in this study. It is likely that several known challenges in the breath VOC field contribute to this^9^. Previous studies have often been structured as pilot studies comparing completely healthy individuals to subjects with established lung cancer. In contrast, the LuCID study focused on an intention-to-diagnose population of subjects with a clinical suspicion of lung cancer, which introduced the heterogeneity necessary for a robust lung cancer diagnostic test. This heterogeneity is exemplified by the substantial overlap between case and control groups, particularly concerning the significantly differentiating VOCs including acetoin and allyl methyl sulphide. The relatively poor signal-to-noise ratio could have several contributing root causes: some VOCs may be (a) general biomarkers of illness, (b) products of common metabolic processes such as oxidative stress, or (c) linked to potential effect modifiers and confounders, such as smoking and age. The substantial size of the LuCID study facilitated the evaluation of these hypotheses.

Another cause for the apparent discrepancy between our results and historical studies likely lies in the statistical toolchains utilised in previous studies. Firstly, many studies were underpowered to adequately interrogate the full VOC-space without significant risk of overfitting. Secondly, multiple testing correction was rarely applied in univariate analyses. Most studies relied solely on cross-validation and did not evaluate the performance of the model on a hold-out set (i.e. validation set) that is completely unexposed to the model training and selection process, as was undertaken in this study. Failure to address these issues and use of split-sample approaches tends to lead to over optimistic p-values and classifier performances.

As a post-hoc analysis, we examined a curated subgroup that excluded subjects with active exacerbations of COPD, infections, or interstitial lung disease. In this analysis, allyl methyl sulphide was found to be significantly lower in breath samples from cases compared to controls, even after multiple testing correction and covariate adjustments (p = 0.044; Table S6), suggesting that this VOC has a better signal-to-noise ratio in this clinically more homogeneous group. However, this analysis did not alter the overall accuracy of the classification model.

In this study we used a GC-MS approach to evaluate the role of VOCs to differentiate lung cancer from co-morbidity matched controls. In recent years several groups have reported on the ability of ‘electronic noses’ to diagnose lung cancer, including in the early stage I, pulmonary nodule setting^17,18,19,20^. Electronic nose technology or ‘E-nose’ utilises a sensor array and machine learning based VOC pattern recognition systems to differentiate between the disease state of interest and controls. Although promising technologies, the challenge for these tools which rely solely on VOC pattern-recognition, is the requirement to meet stringent FDA and EU in-vitro diagnostic regulation analytical validity requirements^21,22^. Regulatory bodies require that devices must demonstrate analytical validity, including calibration, reproducibility, and accuracy. Without known or quantifiable analytes, it is difficult to meet these requirements as standardized measurement and validation are not feasible. Our data demonstrating the significant overlap of VOC profiles between lung cancer and co-morbidity matched controls provides important new insights for the breath research community developing breath biomarkers technologies.

To understand the biological pathways associated with the identified VOCs we performed structural elucidation to obtain definitive chemical identities. Of particular note is Allyl-Methyl-Sulphide, a metabolic end-product of garlic consumption, which was found to be reduced in subjects with lung cancer^23^. While the full biological implications are yet to be elucidated, it is interesting that the effect size of this compound is similar to the magnitude of the known onco-protective effect of garlic consumption^24^. Overall, in this study biological pathway analyses point towards pathways non-specific to lung cancer that are linked to the production of these VOCs. In this context it is of note that many of the literature reported compounds evaluated in this study are hydrocarbons known to be general end-products of oxidative stress^25^.

Given the findings of the LuCID study, it is important to reflect on the potential future of breath analysis for the detection of lung cancer. Careful study design, analytical and data-analysis toolchains and biological interpretation are required for robust biomarker discovery. This underlines the relevance of a staged approach, where biomarker discovery is used to identify VOC-producing pathways for which targeted approaches can be developed to specifically probe these pathways. In line with this, insights from the LuCID study are being used to develop targeted exogenous VOC probing (EVOC-probe) strategies, which evaluate specific pathways associated with lung cancer^26^. This is achieved by administering an exogenous VOC that produces a unique on-breath reporter molecule in response to specific metabolic changes in the tumour. The potential of such approaches to improve the signal-to-noise ratio lies in their ability to generate specific on-breath reporter molecules that are not produced as part of normal metabolism.

In summary, the LuCID study represents the largest breath analysis study for the identification of lung cancer breath biomarkers to date, utilising a very thorough, state-of-the-art toolchain. Although potential breath biomarkers for lung cancer were identified, their diagnostic performance did not surpass that of an established epidemiological risk model. This highlights the necessity for a more targeted approach focusing on pathways associated with lung cancer development to enhance the signal-to-noise ratio in breath analysis.

## Data Availability

Data are available through the corresponding author from the time of publication, following approval of a proposal with a signed data access agreement.

## Acknowledgements

We wish to thank all our patients who participated in the study for their time and help. We are extremely grateful to all the clinical and research staff in participating clinics for patient recruitment and sample collection. Additionally, we wish to acknowledge the contributions of Professor Mina Gaga, Dr Vernoica Conteh, Dr Amrithraj Bhatta, Dr Mark Weatherhead, Dr. Jasper Boschmans, Andrew McLoughlin and Dr John Wrightson (in memoriam).

## Funding

The study was funded by Owlstone Medical Ltd and a UKRI Small Business Research Initiative (SBRI) award. RCR was funded by the NIHR Cambridge Biomedical Research Centre (NIHR203312) and Cancer Research UK Cambridge Centre (CTRQQR-2021\100012).

Conflict of interest statement:

Authors affiliated to Owlstone Medical Ltd were employees and hold options on the company. RCR reports conference funding support from Owlstone Medical Ltd. Owlstone Medical Ltd part-funded research nurses at Royal Papworth Hospital, Cambridge during this work. All remaining authors have declared no conflicts of interest with regard to this work.

## CRediT Authorship Contribution Statement

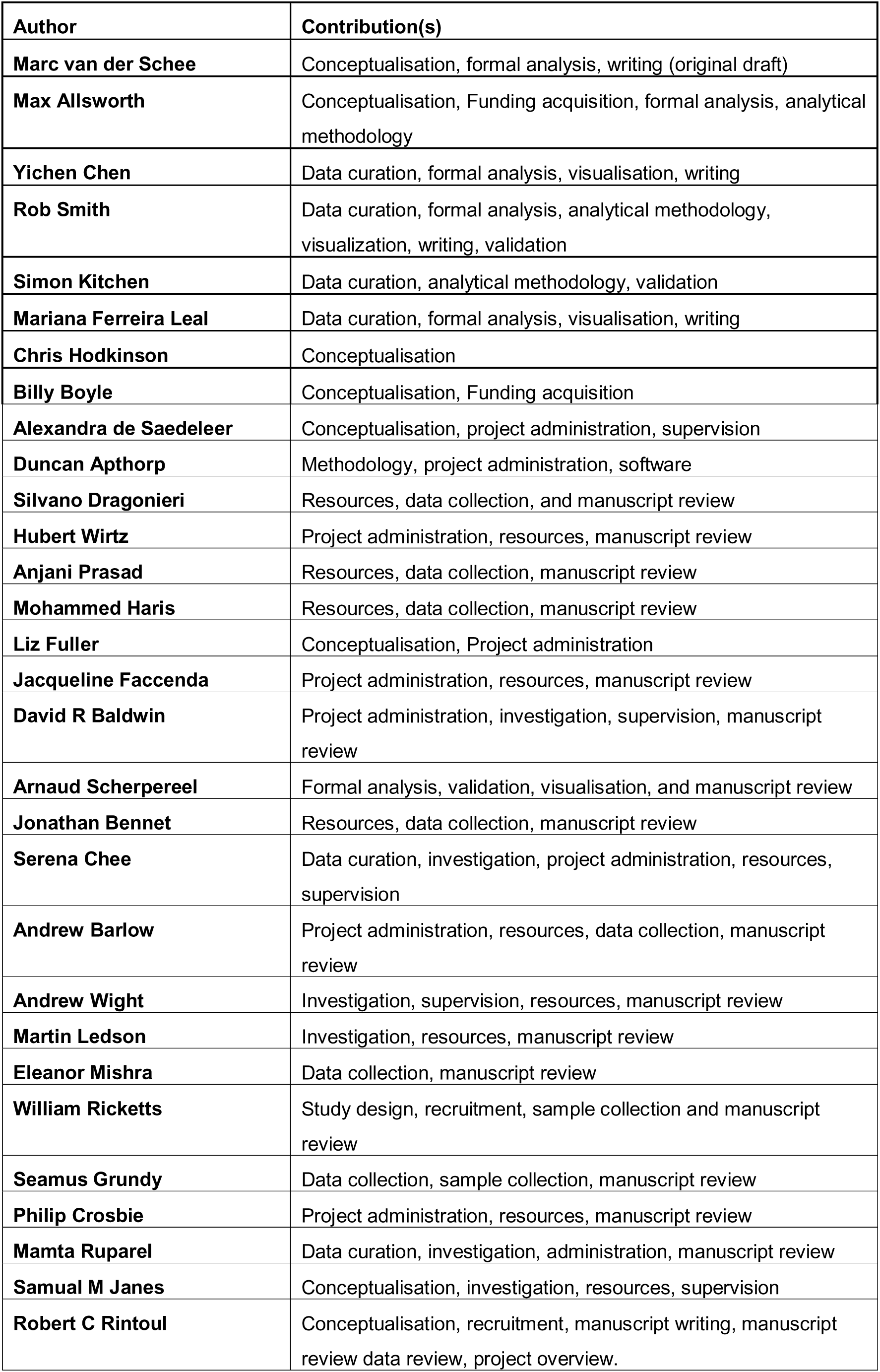

## Notes

### Competing Interest Statement

Authors affiliated to Owlstone Medical were employees and hold options on the company. RCR reports conference funding support from Owlstone Medical. Owlstone Medical Ltd supported research nurse funding at Royal Papworth Hospital, Cambridge during this work. All remaining authors have declared no conflicts of interest with regard to this work.

### Clinical Trial

NCT02612532

### Author Declarations

The study received ethical approval from the National Research Ethics Service Committee East of England; Cambridge South in October 2015 (15/EE/0298).

## References

1 Antoniou SX, Gaude E, Ruparel M, van der Schee MP, Janes SM, Rintoul RC; The LuCID Group. The potential of breath analysis to improve outcome for patients with lung cancer. J Breath Res. 2019 Apr 24;13(3):034002. doi: 10.1088/1752-7163/ab0bee. PMID: 30822771.

2 Wu W, Zhao S. Metabolic changes in cancer: beyond the Warburg effect. Acta Biochim Biophys Sin (Shanghai). 2013 Jan;45(1):18–26. doi: 10.1093/abbs/gms104. PMID: 23257292.

3 Klein EA, Richards D, Cohn A et al. Clinical validation of a targeted methylation-based multi-cancer early detection test using an independent validation set. Ann Oncol. 2021 Sep;32(9):1167–1177. doi: 10.1016/j.annonc.2021.05.806. Epub 2021 Jun 24. PMID: 34176681.

4 de Koning HJ, van der Aalst CM, de Jong PA, et al. Reduced Lung-Cancer Mortality with Volume CT Screening in a Randomized Trial. New England Journal of Medicine. 2020;382(6). doi:10.1056/nejmoa1911793

5 Reduced Lung-Cancer Mortality with Low-Dose Computed Tomographic Screening. New England Journal of Medicine. 2011;365(5):395–409. doi:10.1056/NEJMoa1102873

6 O’Dowd EL, Lee RW, Akram AR et al. Defining the road map to a UK national lung cancer screening programme. Lancet Oncol. 2023 May;24(5):e207–e218. doi: 10.1016/S1470-2045(23)00104-3. PMID: 37142382.

7 Dickson JL, Horst C, Nair A, Tisi S, Prendecki R, Janes SM. Hesitancy around low-dose CT screening for lung cancer. Ann Oncol. 2022; 33: 34–41

8 Out now — Lung cancer screening: learning from implementation - The Lung Cancer Policy Network [Internet]. Available from: https://www.lungcancerpolicynetwork.com/the-lung-cancer-policy-networks-inaugural-report/ (Accessed March 22^nd^ 2025).

9 Tan SY, Ma Q, Li F et al. Does the last 20 years paradigm of clinical research using volatile organic compounds to non-invasively diagnose cancer need to change? Challenges and future direction. J Cancer Res Clin Oncol. 2023 Sep;149(12):10377–10386. doi: 10.1007/s00432-023-04940-7. Epub 2023 Jun 5. PMID: 37273109.

10 Raji OY, Duffy SW, Agbaje OF et al. Predictive accuracy of the Liverpool Lung Project risk model for stratifying patients for computed tomography screening for lung cancer: a case-control and cohort validation study. Ann Intern Med. 2012 Aug 21;157(4):242–50. doi: 10.7326/0003-4819-157-4-201208210-00004. PMID: 22910935; PMCID: PMC3723683.

11 Kitchen S, Edge A, Smith R et al. LATE-BREAKING ABSTRACT: Breathe free: Open-source development of a breath sampler by a consortium of breath researchers. European Respiratory Journal 2015 46: PA3987; doi: 10.1183/13993003.congress-2015.PA3987

12 Horváth I, Barnes PJ, Loukides S et al. A European Respiratory Society technical standard: exhaled biomarkers in lung disease. Eur Respir J. 2017 Apr 26;49(4):1600965. doi: 10.1183/13993003.00965-2016. PMID: 28446552.

13 Wang R, Lu M, An S, Wang J, Yu C. G-Aligner: a graph-based feature alignment method for untargeted LC-MS-based metabolomics. BMC Bioinformatics. 2023 Nov 14;24(1):431. doi: 10.1186/s12859-023-05525-4. PMID: 37964228; PMCID: PMC10644574.

14 Cohen JF, Korevaar DA, Altman DG, Bruns DE, Gatsonis CA, Hooft L, Irwig L, Levine D, Reitsma JB, de Vet HC, Bossuyt PM. STARD 2015 guidelines for reporting diagnostic accuracy studies: explanation and elaboration. BMJ Open. 2016 Nov 14;6(11):e012799. doi: 10.1136/bmjopen-2016-012799. PMID: 28137831; PMCID: PMC5128957.

15 Rudnicka J, Kowalkowski T, Buszewski B. Searching for selected VOCs in human breath samples as potential markers of lung cancer. Lung Cancer. 2019 Sep;135:123–129. doi: 10.1016/j.lungcan.2019.02.012. Epub 2019 Feb 15. PMID: 31446984.

16 Filipiak W, Mochalski P, Filipiak A, et al. A Compendium of Volatile Organic Compounds (VOCs) Released by Human Cell Lines. Curr Med Chem. 2016; 23: 2122–2131

17 Rocco G, Pennazza G, Tan KS, Vanstraelen S, Santonico M, Corba RJ, Park BJ, Sihag S, Bott MJ, Crucitti P, Isbell JM, Ginsberg MS, Weiss H, Incalzi RA, Finamore P, Longo F, Zompanti A, Grasso S, Solomon SB, Vincent A, McKnight A, Cirelli M, Voli C, Kelly S, Merone M, Molena D, Gray K, Huang J, Rusch VW, Bains MS, Downey RJ, Adusumilli PS, Jones DR. A Real-World Assessment of Stage I Lung Cancer Through Electronic Nose Technology. J Thorac Oncol. 2024 Sep;19(9):1272–1283. doi: 10.1016/j.jtho.2024.05.006. Epub 2024 May 16. PMID: 38762120; PMCID: PMC11380592.

18 Kort S, Brusse-Keizer M, Schouwink H, Citgez E, de Jongh FH, van Putten JWG, van den Borne B, Kastelijn EA, Stolz D, Schuurbiers M, van den Heuvel MM, van Geffen WH, van der Palen J. Diagnosing Non-Small Cell Lung Cancer by Exhaled Breath Profiling Using an Electronic Nose: A Multicenter Validation Study. Chest. 2023 Mar;163(3):697–706. doi: 10.1016/j.chest.2022.09.042. Epub 2022 Oct 13. PMID: 36243060.

19 Vadala R, Pattnaik B, Bangaru S, Rai D, Tak J, Kashyap S, Verma U, Yadav G, Dhaliwal RS, Mittal S, Hadda V, Madan K, Guleria R, Agrawal A, Mohan A. A review on electronic nose for diagnosis and monitoring treatment response in lung cancer. J Breath Res. 2023 Mar 27;17(2). doi: 10.1088/1752-7163/acb791. PMID: 36720157.

20 Buma AIG, Muntinghe-Wagenaar MB, van der Noort V, de Vries R, Schuurbiers MMF, Sterk PJ, Schipper S, Meurs J, Cristescu SM, Hiltermann TJN, van den Heuvel MM. Lung cancer detection by electronic nose analysis of exhaled breath: a multi-center prospective external validation study. Ann Oncol. 2025 Mar 31:S0923–7534(25)00125-5. doi: 10.1016/j.annonc.2025.03.013. Epub ahead of print. PMID: 40174676.

21. 21 https://www.ecfr.gov/current/title-21/chapter-I/subchapter-H/part-820 (Accessed 22nd March 2025)

22. 22 https://eur-lex.europa.eu/eli/reg/2017/746/oj/eng (Accessed 22nd March 2025)

23 Temerdashev AZ, Gashimova EM, Porkhanov VA, Polyakov IS, Perunov DV, Dmitrieva EV. Non-Invasive Lung Cancer Diagnostics through Metabolites in Exhaled Breath: Influence of the Disease Variability and Comorbidities. Metabolites. 2023 Jan 30;13(2):203. doi: 10.3390/metabo13020203. PMID: 36837822; PMCID: PMC9960124.

24 Omar SH, Al-Wabel NA. Organosulfur compounds and possible mechanism of garlic in cancer. Saudi Pharm J. 2010 Jan;18(1):51–8. doi: 10.1016/j.jsps.2009.12.007. Epub 2009 Dec 24. PMID: 23960721; PMCID: PMC3731019.

25 Calenic B, Miricescu D, Greabu M et al. “Oxidative stress and volatile organic compounds: interplay in pulmonary, cardio-vascular, digestive tract systems and cancer” Open Chemistry, vol. 13, no. 1, 2015, pp. 000010151520150105. 10.1515/chem-2015-0105

26 Gaude E, Nakhleh MK, Patassini S et al. Targeted breath analysis: exogenous volatile organic compounds (EVOC) as metabolic pathway-specific probes. J Breath Res. 2019 May 17;13(3):032001. doi: 10.1088/1752-7163/ab1789. PMID: 30965287.

