## Supplementary material for "Multi-centre discovery and validation study evaluating breath biomarkers for the diagnosis of lung cancer – the LuCID study": Supplementary data copy.docx

### **Supplemental material**

**Table of contents:**

**List of participating hospitals**

**Inclusion and exclusion criteria**

**Table S1. Challenges limiting translation of breath study results to clinical practice.**

**Table S2. Customized VOC target panel.**

**Table S3. Result of diagnostic work-up for control subjects found not to have lung cancer**

**Table S4. Analysis of VOCs using the *Exploratory* method.**

**Table S5. Analysis of VOCs using the *Optimised* method.**

**Table S6. Analysis of VOCs using the *Optimised* method in subjects without other lung disease.**

**Supplemental data analysis**

**Figure S1. Principal analysis on VOC intensity before and after normalization in the optimized method**

**Figure S2. Elucidation of Unknown 41 breath feature as allyl methyl sulphide on Optimized method**

**Supplemental analytic methods**

**Table S7. Thermal desorption settings for Exploratory and Optimized methods**

**Table S8. GC oven ramp settings for Exploratory method**

**Table S9. GC oven ramp settings for Optimized method**

**Table S10. GC column configuration for Exploratory and Optimized methods**

**Table S11. TOF settings for Exploratory and Optimized methods**

**Table S12. Compounds included in QC mix**

**References**

### **List of participating hospitals**

Università degli Studi di Bari Aldo Morom, Bari, Italy.

Universitätsklinikum Leipzig, Leipzig, Germany.

Wycombe Hospital, Buckinghamshire Healthcare NHS Trust, Wycombe, UK.

Stoke Mandeville Hospital, Buckinghamshire Healthcare NHS Trust, Aylesbury, UK.

Royal Stoke University Hospital, Stoke-on-Trent, UK.

South Tyneside District Hospital, South Shields, UK.

Peterborough City Hospital, Peterborough, UK.

Nottingham University Hospital NHS Trust, Nottingham, UK

Service d'Explorations Fonctionnelles Respiratoires, Hôpital Calmette, Lille, France.

Glenfield Hospital, Leicester, UK.

University Hospital Southampton, Southampton, UK.

West Hertfordshire Teaching Hospitals NHS Trust, Watford, UK.

Arrowe Park Hospital, Wirral, UK.

Liverpool Heart and Chest Hospital NHS Foundation Trust, Liverpool, UK.

Norfolk and Norwich Foundation Trust, Norwich UK

St Bartholomew's Hospital, London, UK.

University Hospital Aintree NHS Foundation Trust, Liverpool, UK.

Manchester University NHS Foundation Trust, Manchester, UK.

University College London, London, UK.

Royal Papworth Hospital, Cambridge, UK (Lead site)

Wansbeck General Hospital, Northumberland, UK

Royal Free Hospital, London, UK

Academisch Medisch Centrum, Amsterdam

Blackpool Teaching Hospitals, Lancashire, UK

Barnet and Chase Farm Hospitals, London, UK

### **Inclusion and exclusion criteria**

Inclusion criteria:

- Older than 18 years at time of consent
- Referred for investigation due to suspicion of lung cancer
  - Referral based on suspicious symptoms
  - Referral based on suspicious finding on imaging, including CT- scan with indeterminate nodule requiring follow-up evaluation.
- Capable of understanding written and/or spoken language
- Able to provide informed consent

Exclusion criteria:

- (Anticipated) inability to complete breath sampling procedure due to

e.g. hyper- or hypo-ventilation, respiratory failure or claustrophobia when wearing the sampling mask

- Participating in a Clinical Trial Investigational Medicinal Product (CTIMP)
- Pulmonary function test with metacholine or beta-2-sympatico mimetic in last 2 hours.
- Any lung biopsy in the past 48 hours
- Currently undergoing anti-cancer treatment for lung cancer

### **Supplemental Tables**

**Table S1. Challenges limiting translation of breath study results to clinical practice.**

| **Issue** | **Consequence** |
| --- | --- |
| **Study design:** design not aligned with intended use population | Inherent population bias not corrected for in analysis |
|  | Performance not assessed relative to current state of the art / additive to established epidemiological risk-model |
| **Analytical Methodology:** Limited attention for quality controls | Pre-analytical; limited attention towards background contamination and consistency of breath collection |
|  | Analytical: No chemical standards run, or technique does not provide any qualitative information resulting in unreliable extraction of candidate biomarkers |
|  | Absence of accurate elucidation of chemicals prohibiting biological validation |
| **Statistical Approach:** Methods used increased risk of overfitted ‘outlier’ results | Global pre-selection approaches use both training and test sets for feature selection |
|  | Small sample size |
|  | No correcting for multiple testing |

**Table S2. Customized VOC target panel.**

| # | **VOC** | **CAS** | **Class of compound** | **Reference** | **Feature included in Exploratory method** | **Feature included in Optimized Method** |
| --- | --- | --- | --- | --- | --- | --- |
| 1 | 1-butanol | 71-36-3 | Alcohol | ^1–4^ | N | Y |
| 2 | 1-Heptanol | 111-70-6 | Alcohol | ^5^ | Y | Y |
| 3 | 1-hexanol | 111-27-3 | Alcohol | ^5^ | Y | Y |
| 4 | 1-Nonanol | 143-08-8 | Alcohol | ^5^ | N | Y |
| 5 | 1-Propanol | 71-23-8 | Alcohol | ^2,5–8^ | Y | N |
| 6 | 2-Butanone | 78-93-3 | Ketone | ^1,2,5,7^ | Y | Y |
| 7 | 2-ethyl-1-hexanol | 104-76-7 | Alcohol | ^2,5,9^ | Y | Y |
| 8 | 2-heptanone | 110-43-0 | Ketone | ^2,5^ | N | Y |
| 9 | 2-heptene | 592-77-8 | Alkene | ^5^ | Y | N |
| 10 | 2-Methylfuran* | 534-22-5 | Furan | ^10^ | Y | Y |
| 11 | 2-methylnonane | 871-83-0 | Alkane | ^11^ | Y | N |
| 12 | 2-methylpentane** | 107-83-5 | Alkane | ^2,5,12^ | Y | Y |
| 13 | 2-methylpropanal | 78-84-2 | Aldehyde | ^2,5,13^ | N | Y |
| 14 | 2-nonanone | 821-55-6 | Ketone | ^2,5^ | Y | Y |
| 15 | 2-pentadecanone | 2345-28-0 | Ketone | ^2,5^ | Y | Y |
| 16 | 2-Pentanone | 107-87-9 | Ketone | ^1,2,4–6,10^ | Y | Y |
| 17 | 2-Propanol | 67-63-0 | Alcohol | ^2,5,10^ | N | N |
| 18 | 2-tridecanone | 593-08-8 | Ketone | ^2,5^ | Y | Y |
| 19 | 2-undecanol | 1653-30-1 | Alcohol | ^5^ | Y | N |
| 20 | 2-undecanone | 112-12-9 | Ketone | ^2,5^ | Y | Y |
| 21 | 3-heptanone | 106-35-4 | Ketone | ^2,5^ | N | Y |
| 22 | 3-hydroxy-2-butanone (acetoin) | 513-86-0 | Acyloins | ^2,3,5,14,15^ | Y | Y |
| 23 | 3-methyl-octane*** | 2216-33-3 | Alkane | ^2,16^ | Y | Y |
| 24 | 3-octanone | 106-68-3 | Ketone | ^5^ | Y | Y |
| 25 | 4-Heptanone | 123-19-3 | Ketone | ^10^ | Y | N |
| 26 | 4-Methyloctane | 3221-61-2 | Alkane | ^5,6,10^ | Y | Y |
| 27 | acetaldehyde | 75-07-0 | Aldehyde | ^2,5,6^ | N | N |
| 28 | Acetone | 67-64-1 | Ketone | ^2,5,8,10^ | Y | Y |
| 29 | Acetophenone | 98-86-2 | Ketone | ^2,5,7,17^ | Y | Y |
| 30 | alpha-pinene | 7785-70-8 | Terpene | ^18^ | Y | Y |
| 31 | b-Pinene | 18172-67-3 | Terpene | ^10^ | Y | Y |
| 32 | Butanal | 123-72-8 | Aldehyde | ^1,2,5,8,13,19^ | N | Y |
| 33 | camphene | 79-92-5 | Terpene | ^20^ | Y | Y |
| 34 | Cyclohexane | 110-82-7 | Alkane | ^1,2,5,10,16^ | N | Y |
| 35 | Cyclohexanone | 108-94-1 | Ketone | ^2,9,10^ | Y | Y |
| 36 | cyclopentane | 287-92-3 | Alkane | ^18^ | Y | N |
| 37 | Decanal | 112-31-2 | Aldehyde | ^1,2^ | Y | Y |
| 38 | Decane | 124-18-5 | Alkane | ^2,12,16^ | Y | Y |
| 39 | Dimethyl sulphide | 75-18-3 | Thioether | ^2,5,6,8,10,18,20^ | Y | Y |
| 40 | Dodecane | 112-40-3 | Alkane | ^1,2,4,10,17^ | Y | Y |
| 41 | Heptanal | 111-71-7 | Aldehyde | ^2,5,19,21^ | Y | Y |
| 42 | Heptane | 142-82-5 | Alkane | ^2,12,20,21^ | Y | Y |
| 43 | hexadecane | 544-76-3 | Alkane | ^2,18^ | Y | Y |
| 44 | Hexanal | 66-25-1 | Aldehyde | ^1,2,4–6,10,16,19–22^ | Y | Y |
| 45 | Hexane | 110-54-3 | Alkane | ^2,4,8,10^ | N | Y |
| 46 | Isobutane | 75-28-5 | Alkane | ^2,10,13^ | N | N |
| 47 | Isoprene | 78-79-5 | Terpene | ^2,4,5,10,12,18^ | Y | Y |
| 48 | Limonene | 5989-54-8 | Terpene | ^10,18,20^ | Y | Y |
| 49 | Malondialdehyde | 542-78-9 | Aldehyde | ^2,23,24^ | N | N |
| 50 | mesitylene | 108-67-8 | Aromatic | ^17^ | Y | Y |
| 51 | Methyl-cyclopentane | 96-37-7 | Alkane | ^2,5,16,17^ | Y | Y |
| 52 | Nonanal | 124-19-6 | Aldehyde | ^2,5,19,20,22^ | Y | Y |
| 53 | Nonane | 111-84-2 | Alkane | ^2,5,6,10,13^ | Y | Y |
| 54 | Octanal | 124-13-0 | Aldehyde | ^2,5,19,20,22^ | Y | Y |
| 55 | octane | 111-65-9 | Alkane | ^2,5,12^ | Y | Y |
| 56 | o-Xylene | 95-47-6 | Aromatic | ^5,6,10,12,18,21^ | Y | Y |
| 57 | p-cresol | 106-44-5 | Aromatic | ^25^ | Y | Y |
| 58 | Pentanal | 110-62-3 | Aldehyde | ^1,2,6,7,13,19,22^ | Y | N |
| 59 | Pentane | 109-66-0 | Alkane | ^2,5,6,12,13^ | N | Y |
| 60 | Propanal | 123-38-6 | Aldehyde | ^1,2,6,8,19^ | N | N |
| 61 | p-Xylene | 106-42-3 | Aromatic | ^2,10,19,26^ | Y | Y |
| 62 | trans-2-hexenol | 928-95-0 | Alcohol/alkene | ^27^ | Y | Y |
| 63 | tridecane | 629-50-5 | Alkane | ^18^ | Y | Y |

*Standard not commercially available; 3-methyl furan used as standard

**Standard not commercially available; 3-methylpentane used as standard

***Standard not commercially available; 2-methyl octane used as standard

**Table S3. Result of diagnostic work-up for control subjects found not to have lung cancer**

| **Diagnosis** | **N** |
| --- | --- |
| No pathology found | 131 |
| COPD exacerbation | 75 |
| Lower respiratory tract infection | 112 |
| Bronchiectasis | 49 |
| Interstitial lung disease | 67 |
| Atelectasis | 34 |
| Hamartoma | 9 |
| Tuberculosis | 5 |
| Amyloidosis | 1 |
| Wegener Granulomatosis | 1 |
| Other benign condition, unspecified | 240 |

**Table S4. Analysis of VOCs using the *Exploratory* method.**

See excel file.

**Table S5. Analysis of VOCs using the *Optimised* method.**

See excel file.

**Table S6. Analysis of VOCs using the *Optimised* method in subjects without other lung disease.**

See excel file.

**Supplemental data-analysis**

Drift in instrument sensitivity over analysis time and across instruments was quantified for each VOC by calculating a running mean of log(intensity/median of all controls) for control subjects over a window of five consecutive sequences. The running mean, once exponentiated, was used as a scaling factor to normalize the intensity of each VOC. The amount of variation associated with analysis time (Figure S1A-B) and instrument (Figure S1C-D) was noticeably reduced after normalization, with the analysis time clusters and instrument clusters no longer present in the first few principal components. Non-analytical variations, such as the differences between current and non-current smokers, were preserved by the normalization process (Figure S1E-F). Both the Optimized method and the Exploratory method exhibited similar changes before and after normalization.

**Power calculation**

Power calculation was based on within-class and between-site variation from the current study by computing the power of a similarly sized study in detecting a single VOC that would have a hypothetical case-control difference equivalent to 60% within-site specificity and sensitivity. The power calculation assumed absence of class imbalance and was based on a single-covariate linear mixed-effect model with a p-value cut-off at 0.05. Further detail is provided in supplementary material.

Within-class standard deviation (free from site effect) and between-site standard deviation were computed from log-transformed VOC intensity and were used as parameters for gaussian distributions from which samples were drawn for simulation. The difference in the means of the case-control gaussian were computed from the required sensitivity and specificity. The standard deviations are unique for each VOC, therefore a whole-panel simulation allows for simultaneous assessment of the power for a range of VOCs characterized by different within-class variation and random site effects.

In each iteration of the simulation, a sample with the same per-class and per-site sample size was randomly drawn. The single-covariate linear mixed effect model (VOC as dependent variable, case-control being the covariate) was fitted to the sample and p-values were calculated. After 500 iterations the estimated power for the Exploratory Method in detecting a single VOC that has a hypothetical within-site effect size equivalent to 60% specificity and 60% sensitivity was at least 99.6%. The estimated power was at least 88.2% for the Optimized Method.

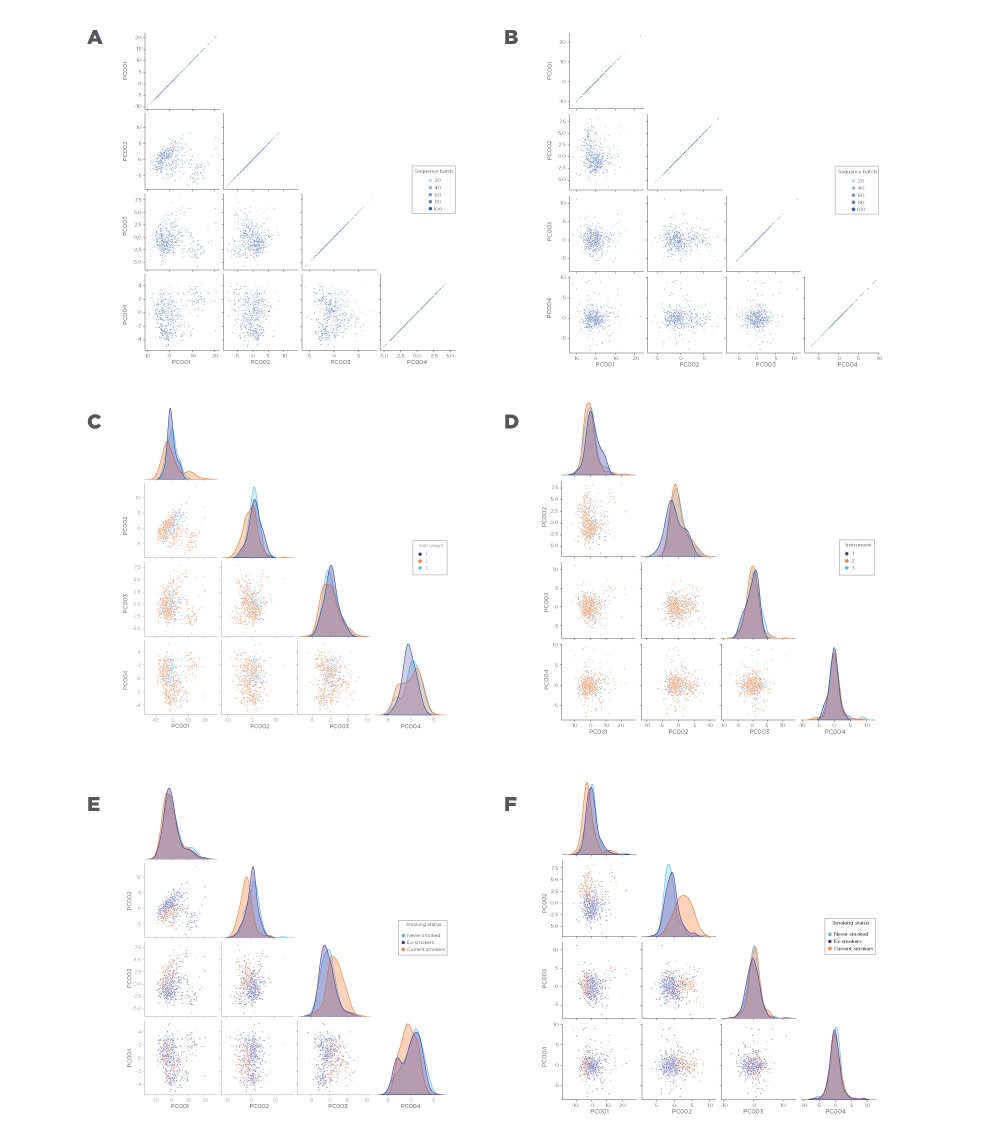

**Figure S1. Principal analysis on VOC intensity before and after normalization in the optimized method.** A) Before normalization by sequence batch; B) After normalization by sequence batch; C) Before normalization by instrument; D) After normalization by instrument; E) Before normalization by subject smoking status; F) After normalization by subject smoking status.

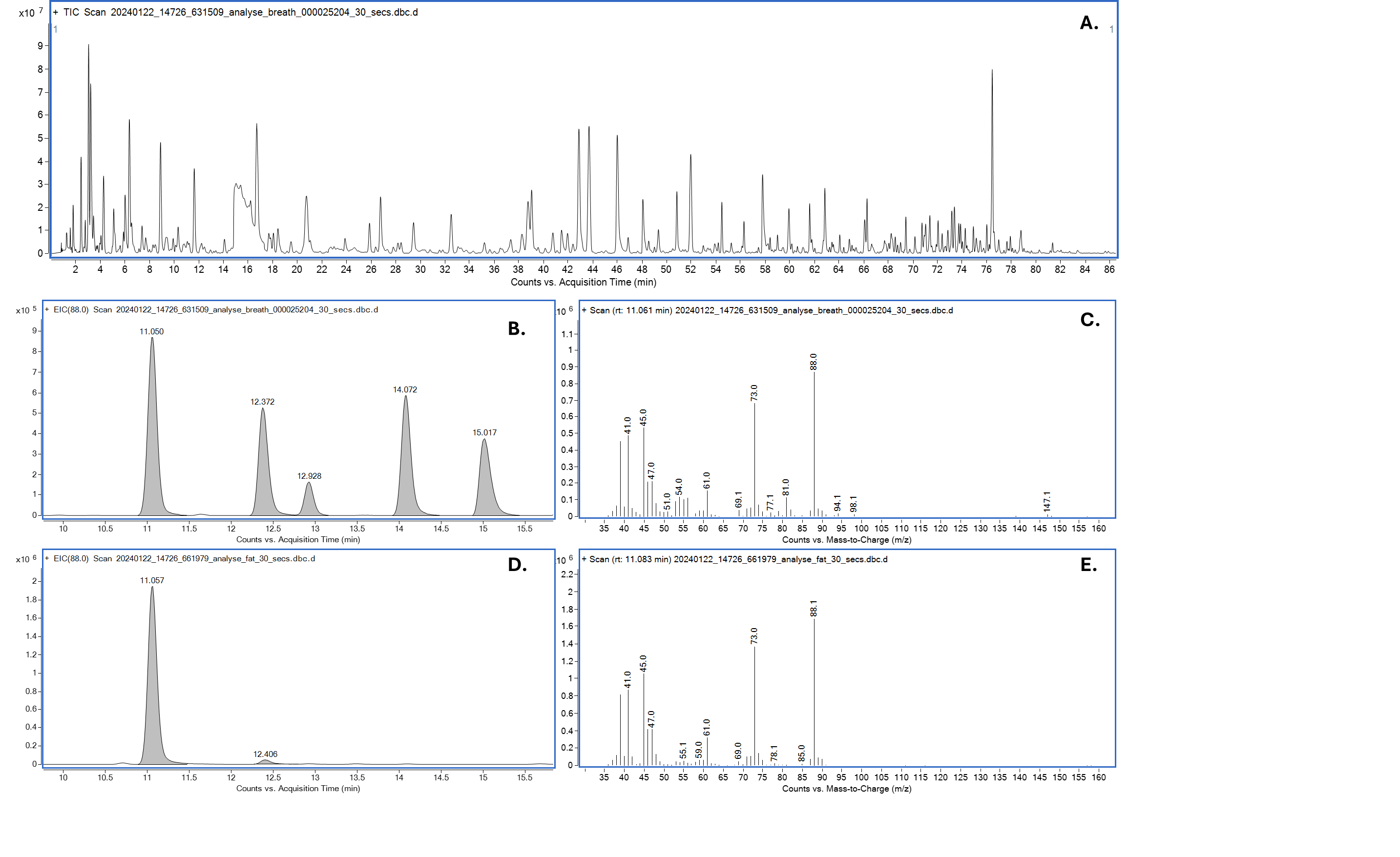

**Figure S2. Elucidation of Unknown 41 breath feature as allyl methyl sulphide on Optimized method,** showing :A. total ion chromatogram of breath sample; B. extracted ion chromatogram (EIC) *m/z* 88 from breath sample; C. mass spectrum of peak at 11.1 mins from breath sample; D. EIC *m/z* 88 of allyl methyl sulphide standard; and E. mass spectrum of peak at 11.1 mins from allyl methyl standard

### **Supplementary analytical methods**

**Elucidation of unknown features**

Unknown VOCs are features that could not be attributed to compounds in the QC mix or the panel of 63 literature-reported compounds. Further elucidation of unknowns aligned with the latest strategies where standard matching with two orthogonal analytical techniques (such as GC retention time and fragmentation mass spectra) provide the highest level of confidence^28,29^. The combination of GC and fragmentation MS were the orthogonal techniques deemed most suitable for elucidation as they were most likely to detect low VOC concentrations on breath at the time of analysis. NIST database searching for compounds with a score exceeding 700 gave a list of candidates, where this score was reduced to 600 if no matches above 700 could be found. The matches were checked based on Kovat's retention index, relative to near eluting known standards with a matched column polarity (low polarity for Exploratory method and medium for Optimized method) to obtain a tentative match. If one or more compounds matched, reference standards of candidates were analysed alongside QC standards and breath samples known to contain the feature being elucidated. A candidate that matches retention time of the feature in breath with close agreement in mass spectra (within 10% of expected relative intensities compared to base peak) will be positively identified as that compound under the standards matched heading and identification post-statistical analysis sub heading of supplementary tables S4, S5a and S5b. Not all features met this criteria; the following resulted in labelling a feature as unknown: i) a feature that does not provide a match with a sufficient NIST ID score (<600); ii) candidates that do not align with breath feature when tested using a standard with no remaining suitable candidates to test (i.e. different elution order relative to other known features); iii) greater than five high scoring NIST matches close to Kovat's retention index, e.g. VOC 23 from the Optimised method was terpenoid in nature where many terpenoids share very similar fragmentation spectra making elucidation very challenging.

**Table S7. Thermal desorption settings for Exploratory and Optimized methods**

|  | Exploratory | Optimized | |
| --- | --- | --- | --- |
| Description | Analysis | Purging | Analysis |
| Type | 2-3 Stage Desorb | 2-3 Stage Desorb | 2-3 Stage Desorb |
| Standby split on | yes | yes | yes |
| Standby flow (mL/min) | 50 | 50 | 50 |
| Flow path temperature (˚C) | 180 | 250 | 250 |
| Split ratio (inlet) | Splitless | - | Splitless |
| Split ratio (outlet) | 2:1 | - | Splitless |
| Split ratio (total) | 2:1 | - | Splitless |
| Pre-purge time (min) | 10 | 12 | - |
| Trap in line | yes | no | - |
| Pre-purge trap flow (mL/min) | 50 | - | - |
| Pre-purge split on | no | yes | - |
| Pre-purge split flow (mL/min) | - | 200 | - |
| Tube desorb time 1 (min) | 5 | - | 10 |
| Tube desorb temp. 1 (˚C) | 300 | - | 210 |
| Trap in line | yes | - | yes |
| Tube desorb trap flow 1 (mL/min) | 50 | - | 50 |
| Tube desorb split on | no | - | no |
| Desorb trap on? | yes | no | yes (*no) |
| Trap purge time (min) | 1 | - | 1 |
| Trap purge flow (mL/min) | 50 | - | 50 |
| Trap low temp. (˚C) | 20 | - | 20 |
| Trap heat rate (˚C/sec) | MAX | - | MAX |
| Trap high temp. (˚C) | 300 | - | 250 |
| Trap desorb time (min) | 3 | - | 3 |
| Trap desorb split on | yes | - | - |
| Trap desorb split flow (mL/min) | 2 | - | - |
| Carrier gas pressure (psi) | 30 | - | 52 |

|  | Exploratory | Optimized | |
| --- | --- | --- | --- |
| Description | Analysis | Purging | Analysis |
| Type | 2-3 Stage Desorb | 2-3 Stage Desorb | 2-3 Stage Desorb |
| Standby split on | yes | yes | yes |
| Standby flow *(mL/min)* | 50 | 50 | 50 |
| Flow path temperature *(˚C)* | 180 | 250 | 250 |
| Split ratio (inlet) | Splitless | - | Splitless |
| Split ratio (outlet) | 2:1 | - | Splitless |
| Split ratio (total) | 2:1 | - | Splitless |
| Pre-purge time *(min)* | 10 | 12 | - |
| Trap in line | yes | no | - |
| Pre-purge trap flow *(mL/min)* | 50 | - | - |
| Pre-purge split on | no | yes | - |
| Pre-purge split flow (mL/min) | - | 200 | - |
| Tube desorb time 1 *(min)* | 5 | - | 10 |
| Tube desorb temp. 1 *(˚C)* | 300 | - | 210 |
| Trap in line | yes | - | yes |
| Tube desorb trap flow 1 *(mL/min)* | 50 | - | 50 |
| Tube desorb split on | no | - | no |
| Desorb trap on? | yes | no | yes (*no) |
| Trap purge time *(min)* | 1 | - | 1 |
| Trap purge flow *(mL/min)* | 50 | - | 50 |
| Trap low temp. *(˚C)* | 20 | - | 20 |
| Trap heat rate *(˚C/sec)* | MAX | - | MAX |
| Trap high temp. *(˚C)* | 300 | - | 250 |
| Trap desorb time *(min)* | 3 | - | 3 |
| Trap desorb split on | yes | - | - |
| Trap desorb split flow *(mL/min)* | 2 | - | - |
| Carrier gas pressure (psi) | 30 | - | 52 |

*First sample tube out of pair of tubes run without cold trap heating. Cold trap desorbed both first and second sample tubes to provide a 2-fold increase in amount of sample loaded in a single run.

**Table S8. GC oven ramp settings for Exploratory method**

| **Step** | **Rate (˚C/min.)** | **Temp. (˚C)** | **Hold time (min.)** |
| --- | --- | --- | --- |
| Initial | - | 50.0 | 4.50 |
| 1 | 2.5 | 100.0 | 0.00 |
| 2 | 7.5 | 160.0 | 0.00 |
| 3 | 25.0 | 310.0 | 8.00 |
| 4 | 50.0 | 50.0 | 8.50 |

**Table S9. GC oven ramp settings for Optimized method**

| **Step** | **Rate (˚C/min.)** | **Temp. (˚C)** | **Hold time (min.)** |
| --- | --- | --- | --- |
| Initial | - | 40 | 4.00 |
| 1 | 1.50 | 100 | 0.00 |
| 2 | 4.00 | 250 | 5.00 |

**Table S10. GC column configuration for Exploratory and Optimized methods**

| **Description** | **Exploratory method** | **Optimized method** |
| --- | --- | --- |
| Column type | VF-5ms | Quadrex 007-624 |
| Column length | 60 m | 30 m |
| Column diam. | 0.250 mm | 0.32 mm |
| Column film thickness | 0.250 μm | 3 µm |
| Pressure or flow controlled | Pressure | Flow |
| Column flow | 1.94 mL/min. | 3.0 mL/min |

**Table S11. TOF settings for Exploratory and Optimized methods**

| **Description** | **Exploratory** | **Optimized** |
| --- | --- | --- |
| Run time (mins) | 59 | 86.5 |
| Filament voltage *(V)* | 1.5 | 1.6 |
| Filament delay *(sec)* | 6 | 6 |
| Filament standby voltage *(V)* | 1 | 1 |
| Timed events (time) | 0.10 / 5.30 / 39.00 | 0.10 |
| Timed events (voltage) | 1.50 / 1.70 / 1.50 | 1.6 |
| Start mass *(m/z)* | 35.0 | 35.0 |
| End mass *(m/z)* | 350.0 | 350.0 |
| Data rate (scans/sec) | 1.500 | 1.500 |
| Transfer line temp. *(˚C)* | 310 | 310 |
| Ion source temp. *(˚C)* | 275 | 310 |
| Ionisation energy *(eV)* | -70.000 | -70.000 |

**Table S12. Compounds included in QC mix**

| Compound | CAS |
| --- | --- |
| 1,2-Dichlorobenzene | 95-50-1 |
| 1,2-Dichloropropane | 78-87-5 |
| 1-Decene | 872-05-9 |
| 1-Dodecene | 112-41-4 |
| 1-Heptene | 592-76-7 |
| 1-Hexene | 592-41-6 |
| 1-Nonene | 124-11-8 |
| 1-Octene | 111-66-0 |
| 1-Pentene | 109-67-1 |
| 1-Tetradecene | 1120-36-1 |
| 1-Undecene | 821-95-4 |
| 2-Butanone | 78-98-3 |
| 2-Pentanone | 107-87-9 |
| Acetone | 67-64-1 |
| Butanal | 123-72-8 |
| Chlorobenzene | 108-90-7 |
| cis-1,2-Dichloroethene | 156-59-2 |
| Decane | 124-18-5 |
| D-Limonene | 5989-27-5 |
| Dodecane | 112-40-3 |
| Ethylbenzene | 100-41-4 |
| Heptane | 142-82-5 |
| Hexane | 110-54-3 |
| Isoprene | 78-79-5 |
| Nonane | 111-84-2 |
| Octane | 111-65-9 |
| o-Xylene | 95-47-6 |
| Pentadecane | 629-62-9 |
| Pentane | 109-66-0 |
| p-Xylene | 106-42-3 |
| Styrene | 100-42-5 |
| Tetrachloroethene | 127-18-4 |
| Tetradecane | 629-59-4 |
| Toluene | 108-88-3 |
| Tridecane | 629-50-5 |
| Undecane | 1120-21-4 |

### **References**

1. Schallschmidt K, Becker R, Jung C, et al. Comparison of volatile organic compounds from lung cancer patients and healthy controls—challenges and limitations of an observational study. J Breath Res 2016; 10: 046007.
2. COD Cancer Odor Database. http://bioinf.modares.ac.ir/software/COD/index.html (accessed Sept 9, 2024).
3. Song G, Qin T, Liu H, et al. Quantitative breath analysis of volatile organic compounds of lung cancer patients. Lung Cancer 2010; 67: 227–31.
4. Gashimova E, Osipova A, Temerdashev A, et al. Exhaled breath analysis using GC-MS and an electronic nose for lung cancer diagnostics. Anal Methods 2021; 13: 4793–804.
5. Filipiak W, Mochalski P, Filipiak A, et al. A Compendium of Volatile Organic Compounds (VOCs) Released By Human Cell Lines. CMC 2016; 23: 2112–31.
6. Ulanowska A, Kowalkowski T, Trawińska E, Buszewski B. The application of statistical methods using VOCs to identify patients with lung cancer. J Breath Res 2011; 5: 046008.
7. Bajtarevic A, Ager C, Pienz M, et al. Noninvasive detection of lung cancer by analysis of exhaled breath. BMC Cancer 2009; 9: 348.
8. Kischkel S, Miekisch W, Sawacki A, et al. Breath biomarkers for lung cancer detection and assessment of smoking related effects — confounding variables, influence of normalization and statistical algorithms. Clinica Chimica Acta 2010; 411: 1637–44.
9. Liu H, Wang H, Li C, Wang L, Pan Z, Wang L. Investigation of volatile organic metabolites in lung cancer pleural effusions by solid-phase microextraction and gas chromatography/mass spectrometry. Journal of Chromatography B 2014; 945–946: 53–9.
10. Rudnicka J, Kowalkowski T, Buszewski B. Searching for selected VOCs in human breath samples as potential markers of lung cancer. Lung Cancer 2019; 135: 123–9.
11. Filipiak W, Ruzsanyi V, Mochalski P, et al. Dependence of exhaled breath composition on exogenous factors, smoking habits and exposure to air pollutants. J Breath Res 2012; 6: 036008.
12. Poli D, Carbognani P, Corradi M, et al. Exhaled volatile organic compounds in patients with non-small cell lung cancer: cross sectional and nested short-term follow-up study. Respir Res 2005; 6: 71.
13. Rudnicka J, Kowalkowski T, Ligor T, Buszewski B. Determination of volatile organic compounds as biomarkers of lung cancer by SPME–GC–TOF/MS and chemometrics. Journal of Chromatography B 2011; 879: 3360–6.
14. Fu X, Li M, Knipp RJ, Nantz MH, Bousamra M. Noninvasive detection of lung cancer using exhaled breath. Cancer Medicine 2014; 3: 174–81.
15. Smirnova E, Mallow C, Muschelli J, et al. Predictive performance of selected breath volatile organic carbon compounds in stage 1 lung cancer. Transl Lung Cancer Res 2022; 11: 1009–18.
16. Phillips M, Gleeson K, Hughes JMB, et al. Volatile organic compounds in breath as markers of lung cancer: a cross-sectional study. The Lancet 1999; 353: 1930–3.
17. Cheng X, Feng Y, Chen S, et al. Rapid Point-of-Care Exhaled Breath Analysis for Lung Cancer Diagnosis Using a Micro Gas Chromatography System: A Pilot Study. 2024; published online June 27. DOI:10.1101/2024.06.27.24309565.
18. Choueiry F, Barham A, Zhu J. Analyses of lung cancer-derived volatiles in exhaled breath and in vitro models. Exp Biol Med (Maywood) 2022; 247: 1179–90.
19. Poli D, Goldoni M, Corradi M, et al. Determination of aldehydes in exhaled breath of patients with lung cancer by means of on-fiber-derivatisation SPME–GC/MS. Journal of Chromatography B 2010; 878: 2643–51.
20. Zou Y, Hu Y, Jiang Z, et al. Exhaled metabolic markers and relevant dysregulated pathways of lung cancer: a pilot study. Annals of Medicine 2022; 54: 790–802.
21. Chen X, Muhammad KG, Madeeha C, et al. Calculated indices of volatile organic compounds (VOCs) in exhalation for lung cancer screening and early detection. Lung Cancer 2021; 154: 197–205.
22. Fuchs P, Loeseken C, Schubert JK, Miekisch W. Breath gas aldehydes as biomarkers of lung cancer. Intl Journal of Cancer 2010; 126: 2663–70.
23. Gonenc A, Ozkan Y, Torun M, Simsek B. Plasma malondialdehyde (MDA) levels in breast and lung cancer patients. J Clin Pharm Ther 2001; 26: 141–4.
24. Peddireddy V, Siva Prasad B, Gundimeda SD, Penagaluru PR, Mundluru HP. Assessment of 8-oxo-7, 8-dihydro-2′-deoxyguanosine and malondialdehyde levels as oxidative stress markers and antioxidant status in non-small cell lung cancer. Biomarkers 2012; 17: 261–8.
25. Peralbo-Molina A, Calderón-Santiago M, Priego-Capote F, Jurado-Gámez B, Luque De Castro MD. Identification of metabolomics panels for potential lung cancer screening by analysis of exhaled breath condensate. J Breath Res 2016; 10: 026002.
26. Koureas M, Kalompatsios D, Amoutzias GD, Hadjichristodoulou C, Gourgoulianis K, Tsakalof A. Comparison of Targeted and Untargeted Approaches in Breath Analysis for the Discrimination of Lung Cancer from Benign Pulmonary Diseases and Healthy Persons. Molecules 2021; 26: 2609.
27. Furuhashi T, Matsumoto Y, Ishii R, Sugasawa T, Ota S. Hypoxia and lactate influence VOC production in A549 lung cancer cells. Front Mol Biosci 2023; 10: 1274298.
28. Raspotnig G, Rohlfs M. A matter of confidence: requirements and standards for compound identification in Chemoecology. Chemoecology. 2023; 33; 145-146
29. Schymanski EL, Jeon J,  Gulde R *et al.* Identifying Small Molecules via High Resolution Mass Spectrometry: Communicating Confidence. Environ. Sci. Technol. 2014; 48, 2097-2098
